# Critical Illness Risk and Long-Term Outcomes Following Intensive Care in Pediatric Hematopoietic Cell Transplant Recipients

**DOI:** 10.1101/2023.07.31.23293444

**Authors:** Matt S. Zinter, Ruta Brazauskas, Joelle Strom, Stella Chen, Stephanie Bo-Subait, Akshay Sharma, Amer Beitinjaneh, Dimana Dimitrova, Greg Guilcher, Jaime Preussler, Kasiani Myers, Neel S. Bhatt, Olle Ringden, Peiman Hematti, Robert J. Hayashi, Sagar Patel, Satiro Nakamura De Oliveira, Seth Rotz, Sherif M. Badawy, Taiga Nishihori, David Buchbinder, Betty Hamilton, Bipin Savani, Hélène Schoemans, Mohamed Sorror, Lena Winestone, Christine Duncan, Rachel Phelan, Christopher C. Dvorak

**Affiliations:** Department of Pediatrics, Division of Critical Care Medicine, University of California, San Francisco, San Francisco, CA, USA; Department of Pediatrics, Division of Allergy, Immunology, and BMT, University of California, San Francisco, San Francisco, CA, USA; Medical College of Wisconsin, Milwaukee, WI, USA; St. Jude Children’s Research Hospital, Memphis, TN, USA; University of Miami, Miami, FL, USA; National Institutes of Health, National Cancer Institute, Bethesda, MD, USA; Alberta Children’s Hospital, Alberta, Canada; National Marrow Donor Program/Be The Match, Minneapolis, MN, USA; Cincinnati Children’s Hospital Medical Center, Cincinnati, OH, USA; Fred Hutchinson Cancer Center, Seattle, WA, USA; Karolinska Institutet, Karolinska University Hospital, Huddinge, The Netherlands; Washington University in St. Louis, St. Louis, MO, USA; Huntsman Cancer Institute, University of Utah, Salt Lake City, UT, USA; University of California, Los Angeles, Los Angeles, CA, USA; Cleveland Clinic, Cleveland, OH, USA; Northwestern University Feinberg School of Medicine, Chicago, IL, USA; Moffitt Cancer Center, Tampa, FL, USA; Children’s Hospital of Orange County, Orange, CA, USA; Vanderbilt University Medical Center, Nashville, TN, USA; University Hospital Gasthuisberg, Leuven, Belgium; Dana Farber Cancer Center, Boston, MA, USA

## Abstract

**Background:** Allogeneic hematopoietic cell transplantation (HCT) can be complicated by the development of organ toxicity and infection necessitating intensive care. Risk factors for intensive care admission are unclear due to heterogeneity across centers, and long-term outcome data after intensive care are sparse due to a historical paucity of survivors.

**Methods:** The Center for International Blood and Marrow Transplant Research (CIBMTR) was queried to identify patients age ≤21 years who underwent a 1^st^ allogeneic HCT between 2008-2014 in the United States or Canada. Records were cross-referenced with the Virtual Pediatric Systems pediatric ICU database to identify intensive care admissions. CIBMTR follow-up data were collected through the year 2020.

**Results:** We identified 6,995 pediatric HCT patients from 69 HCT centers, of whom 1,067 required post-HCT intensive care. The cumulative incidence of PICU admission was 8.3% at day +100, 12.8% at 1 year, and 15.3% at 5 years post HCT. PICU admission was linked to younger age, lower median zip code income, Black or multiracial background, pre-transplant organ toxicity, pre-transplant CMV seropositivity, use of umbilical cord blood and/or HLA-mismatched allografts, and the development of post-HCT graft-versus-host disease or malignancy relapse. Among PICU patients, survival to ICU discharge was 85.7% but more than half of ICU survivors were readmitted to a PICU during the study interval. Overall survival from the time of 1^st^ PICU admission was 52.5% at 1 year and 42.6% at 5 years. Long-term post-ICU survival was worse among patients with malignant disease (particularly if relapsed), as well as those with poor pre-transplant organ function and alloreactivity risk-factors. In a landmark analysis of all 1-year HCT survivors, those who required intensive care in the first year had 10% lower survival at 5 years (77.1% vs. 87.0%, p<0.001) and developed new dialysis-dependent renal failure at a greater rate (p<0.001).

**Conclusions:** Intensive care management is common in pediatric HCT patients. Survival to ICU discharge is high, but ongoing complications necessitate recurrent ICU admission and lead to a poor 1-year outcome in many patients. Together, these data suggest an ongoing burden of toxicity in pediatric HCT patients that continues to limit long-term survival.

## BACKGROUND

Allogeneic hematopoietic cell transplantation (HCT) offers a potential cure to over thousands of children annually across the world, including those with high-risk leukemia, immunodeficiencies, hemoglobinopathies, and other life threatening disorders.^1^ However, a key barrier to HCT success is the development of acute organ failure due to chemotherapeutic toxicity, radiation exposure, infection, genetic predisposition, and impaired or dysregulated immunity.^2, 3^ Children requiring intensive care unit (ICU) admission suffer >20% mortality, with rates exceeding 45% when intubation and mechanical ventilation are required.^4–6^

A prominent goal of transplant and intensive care physicians is to quickly and correctly identify high-risk patients to stop the advancement of critical illness and prevent irreversible organ failure.^7^ This is predicated on compelling evidence indicating that organ failure can be modified by promptly recognizing and intervening in its early stages.^8–11^ However, predicting who will require intensive care after HCT has been challenging due to relatively small patient numbers spread out across multiple institutions with varying intensive care unit (ICU) admission criteria. Specifically, HCT databases do not capture ICU transfer data and ICU databases do not rigorously phenotype HCT complexity, precluding deeper analyses of risk for critical illness.^12^

All major international HCT organizations recommend following all pediatric patients who undergo HCT for long-term cardiopulmonary, renal, and multiorgan toxicities as well as for neurodevelopmental outcomes and health-related quality of life.^13–15^ These recommendations are even more crucial in light of estimates that the number of pediatric survivors of HCT doubled between 2009 and 2020, and will likely double again between 2020 and 2030 to reach an estimated 64,000 patients. However, while recent reports have shown improved survival to ICU discharge, long-term outcomes are lacking since ICU databases do not typically follow patients past ICU discharge.^4–6, 16^ In one report of pediatric HCT patients who survived to pediatric ICU (PICU) discharge, survival at 1 year was 40% relative to 1-year survival of 65% in patients who never required PICU admission; this suggests an ongoing burden of chronic organ toxicity in these vulnerable patients.^17^ Taken together, these data suggest that both short- and long-term outcomes for critically ill pediatric HCT patients remain suboptimal, and that the toxicity of HCT remains a major barrier to a disease-free childhood for many patients.^18, 19^

To address ongoing knowledge gaps regarding risk-factors for critical illness and long-term outcomes, we merged records from two large research databases to determine the incidence of and risk-factors for post-HCT intensive care. We followed patients requiring PICU admission for a median of 5-years to determine long-term outcomes. To assess the burden of chronic toxicities, we also compared survival and organ dysfunction in 1-year transplant survivors according to a preceding need for intensive care in the first year.

## METHODS

### Data Sources

The Center For International Blood and Marrow Transplant Research (CIBMTR) is a research collaboration between the National Marrow Donor Program®/Be The Match® and the Medical College of Wisconsin. It comprises over 450 transplant centers worldwide that contribute high-quality longitudinal data on consecutive allogeneic HCT patients. Participating centers contribute data about individual patients and their exposures and outcomes. CIBMTR data is collected and reported at two levels: Transplant Essential Data (TED) and Comprehensive Report Form (CRF). TED data include disease type, age, sex, pretransplant disease stage, and chemotherapy responsiveness, date of diagnosis, graft type, conditioning regimen, posttransplant disease progression and survival, development of a new malignancy, and cause of death. All CIBMTR centers contribute TED data. More detailed clinical information is collected via the CRF mechanism for a subset of randomly selected patients. Observational studies conducted by the CIBMTR are performed in compliance with all applicable federal regulations pertaining to the protection of human research participants. The Virtual Pediatric Systems (VPS) database documents PICU admissions across over 140 pediatric hospitals predominantly in the United States and Canada. Admission characteristics, severity of illness scores, critical care interventions, and critical care-related diagnosis codes are documented by trained analysts at each site, with >95% inter-rater reliability. Patients are followed until hospital discharge.

The CIBMTR database was queried for patients who underwent first allogeneic HCT between 2008-2014 at an age ≤21 years in the United States or Canada and were at a HCT site that also submitted data to VPS. Patients were excluded for allograft source other than bone marrow, peripheral blood, or umbilical cord blood; syngeneic donor; lack of consent to use data; or lack of ≥100 days follow-up. Use of intensive care at any point after first allogeneic HCT was identified by matching records between CIBMTR and VPS databases as previously described.^4^ To avoid analyzing low-risk patients, intensive care admissions designated as perioperative or scheduled (>12 hours notice) lasting <2 days were excluded. All PICU admission data were benchmarked to date of first allogeneic HCT since data on repeat allogeneic HCT could not be reliably differentiated from other cellular therapies (eg: donor lymphocyte infusion) in the CIBMTR database during this period of data collection. Patients admitted to an adult ICU were not included. Follow-up data were reported to CIBMTR and were abstracted for this study in the first quarter of 2020.

### Outcomes

The primary outcomes were the need for intensive care and long-term mortality after intensive care. We performed a landmark analysis of all patients alive at transplant day +365 to determine if prior need for intensive care was associated with long-term morbidity and mortality. Approximately half of CIBMTR patients participated in the data-intensive CRF track; for these patients, the occurrence and start date of the following toxicities were collected: congestive heart failure; non-infectious pulmonary dysfunction (interstitial pneumonitis and other non-infectious pulmonary abnormalities including bronchiolitis obliterans, COP/BOOP, and diffuse alveolar hemorrhage); renal failure severe enough to warrant dialysis; non-infectious liver toxicity (including sinusoidal obstruction syndrome, cirrhosis, and other); stroke/seizure; and diabetes/hyperglycemia. Clinical changes in each toxicity, such as worsening, improvement, or resolution, were not captured in CIBMTR.

### Covariates

We considered variables in the following categories. ***Demographics***: age at HCT, sex, race, ethnicity, insurance status, zip code median income, weight classification at HCT. ***Pre-HCT:*** disease, disease status prior to HCT (for malignancy only), HCT Comorbidity Index (HCT-CI), pre-HCT Lansky/Karnofsky, history of pre-HCT mechanical ventilation, history of pre-HCT invasive fungal infection. ***HCT-related:*** HCT center size, time from diagnosis to HCT (for malignancy only), graft source, donor type/match, allograft manipulation, donor/recipient blood types, donor/recipient CMV status, donor/recipient sex matching, conditioning regimen intensity, conditioning regimen serotherapy, GVHD prophylaxis regimen. ***Post-HCT:*** achievement of neutrophil engraftment, aGVHD grade, cGVHD grade, hematologic malignancy relapse. ***ICU-related:*** PICU center size (according to tertile of HCT patient PICU admission volume), age at PICU admission, weight category at PICU admission, time interval between HCT and at PICU admission, Pediatric Risk of Mortality Score-3 (PRISM-3); use of invasive mechanical ventilation (IMV), non-invasive mechanical ventilation (NIV), or renal replacement therapy (RRT); and presence of Gram-positive, Gram-negative, viral, or fungal infection. The PRISM-3 score ranges from 0 to 47 and is composed of 17 vital sign and laboratory derangements measured in the first 12 hours of PICU admission.^20^

### Statistical Approach

Descriptive statistics were used to summarize patient characteristics. Cumulative incidence of being admitted to the intensive care unit was calculated with death being treated as competing risk. Cox proportional hazard model was used to identify factors associated with the need for intensive care. Stepwise model selection was used to identify predictors significantly associated with the outcome. Only variables significant at the 0.05 level were retained in the final model. Overall survival probabilities after intensive care admission were calculated using the Kaplan-Meier estimate. Cox proportional hazard regression with stepwise model selection was used to identify risk factor associated with mortality among patients who were admitted to PICU. Covariates which were not known at baseline (e.g., aGVHD, cGVHD, relapse, use of invasive and non-invasive ventilation) were treated as time-dependent covariates. Next, we explored the difference in survival between patients who were admitted to PICU within 1 year after HCT and those who were not. Only patients surviving to Day +365 post HCT were included in this analysis. Survival after day +365 was estimated using Kaplan Meier curves and factors associated with overall mortality were assessed using Cox proportional hazard regression with stepwise model selection. The development of post-HCT organ toxicities among patients alive at day +365 was assessed in the CRF-track subset of patients using the cumulative incidence function. Nominal significance was considered at a p value <0.05.

## RESULTS

### Patients

The final cohort included 6,995 pediatric HCT patients from 69 HCT centers; 1,067 patients required critical care across 79 PICUs (**Figure 1a**). Baseline characteristics of included patients are listed in **Table 1** and **Supplemental Table 1**. The underlying reason for allogeneic HCT was malignancy in 57%. Myeloablative conditioning chemotherapy was used in 73% of patients, including total body irradiation (TBI) in 35% of patients. HCT allografts were fully HLA-matched in 54% of patients and obtained most commonly from harvested bone marrow (57%). The largest third of HCT centers (n=23 out of 69) accounted for 67% of all 1^st^ allogeneic transplants performed.

**Figure 1:**
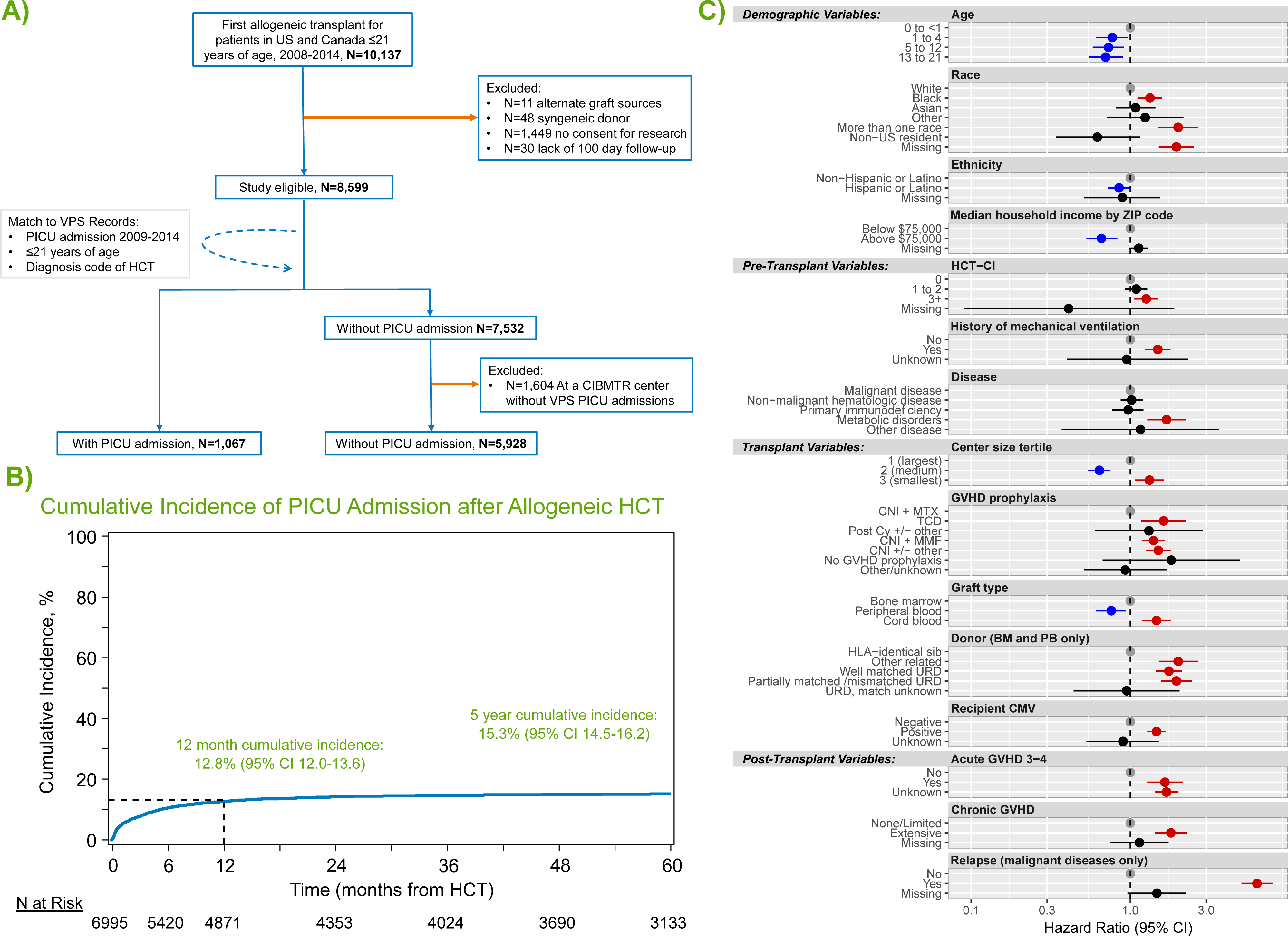
(A) Inclusion/exclusion flow diagram. (B) Cumulative incidence of PICU admission after allogeneic HCT. (B) Factors independently associated with post-HCT PICU admission in multivariable competing risk-regression model.

### Need for Intensive Care

Following HCT, the cumulative incidence of PICU admission was 8.3% at day +100 (95% CI 7.7-9.0), 12.8% at 1 year (95% CI 12.0-13.6), and 15.3% at 5 years post HCT (95% CI 14.5-16.2, **Figure 1b**).

Multivariable analysis with stepwise variable selection identified 15 risk-factors independently associated with the need for intensive care (**Figure 1c; Supplemental Table 2**). Among demographic factors, younger age, lower median ZIP code income, Black race (HR 1.33, 95% CI 1.11-1.59, p=0.002), multi-racial background (HR 2.0, 95% CI 1.50-2.67, p<0.001), and non-Hispanic ethnicity were each associated with PICU admission. Pre-transplant organ toxicity was also associated with greater rates of PICU admission, as measured by an HCT-CI score of ≥3 (HR 1.26, 95% CI 1.06-1.49, p=0.008) or a history of mechanical ventilation (HR 1.49, 95% CI 1.24-1.79, p<0.001). Several transplant-related variables were associated with PICU admission, including transplantation at centers in the lowest tertile of transplant volume (HR 1.32, 95% CI 1.07-1.63, p=0.009), HCT for underlying inborn errors of metabolism (HR 1.69, 95% CI 1.28-2.23, p<0.001), and recipient CMV positivity (HR 1.46, 95% CI 1.28-1.67, p<0.001). PICU admission was also associated with the use of umbilical cord blood allografts (HR 1.46, 95% CI 1.18-1.81, p<0.001) and donors other than HLA-identical siblings (eg: partially matched unrelated donor HR 1.95, 95% CI 1.57-2.43, p<0.001). Finally, the development of post-transplant complications was strongly associated with subsequent PICU admission, including aGVHD grade 3-4 (HR 1.65, 95% CI 1.28-2.14, p<0.001), extensive cGVHD (HR 1.80, 95% CI 1.43-2.28, p<0.001), and post-HCT relapse of malignancy (HR 6.26, 95% CI 5.00-7.84, p<0.001).

### Outcomes after Intensive Care Admission

Characteristics of patients at the time of 1^st^ PICU admission are listed in **Table 2**. The per-ICU admission survival rate was 85.7% with a median ICU LOS of 3 days (IQR 1-11). Whereas 14% died in the first ICU admission, 44% survived but required at least one additional PICU admission and 42% of patients survived and did not require PICU re-admission during the study interval. Using CIBMTR longitudinal data, patients were followed from the time of ICU admission to a median 73 months (range 3-147). The OS from the time of 1^st^ PICU admission was 52.5% at 1 year (95% CI 49.5-55.5%), 44.4% at 3 years (95% CI 41.5-47.5%), and 42.6% at 5 years (95% CI 39.6-45.6%, **Figure 2a, Supplemental Tables 3-4**).

**Figure 2:**
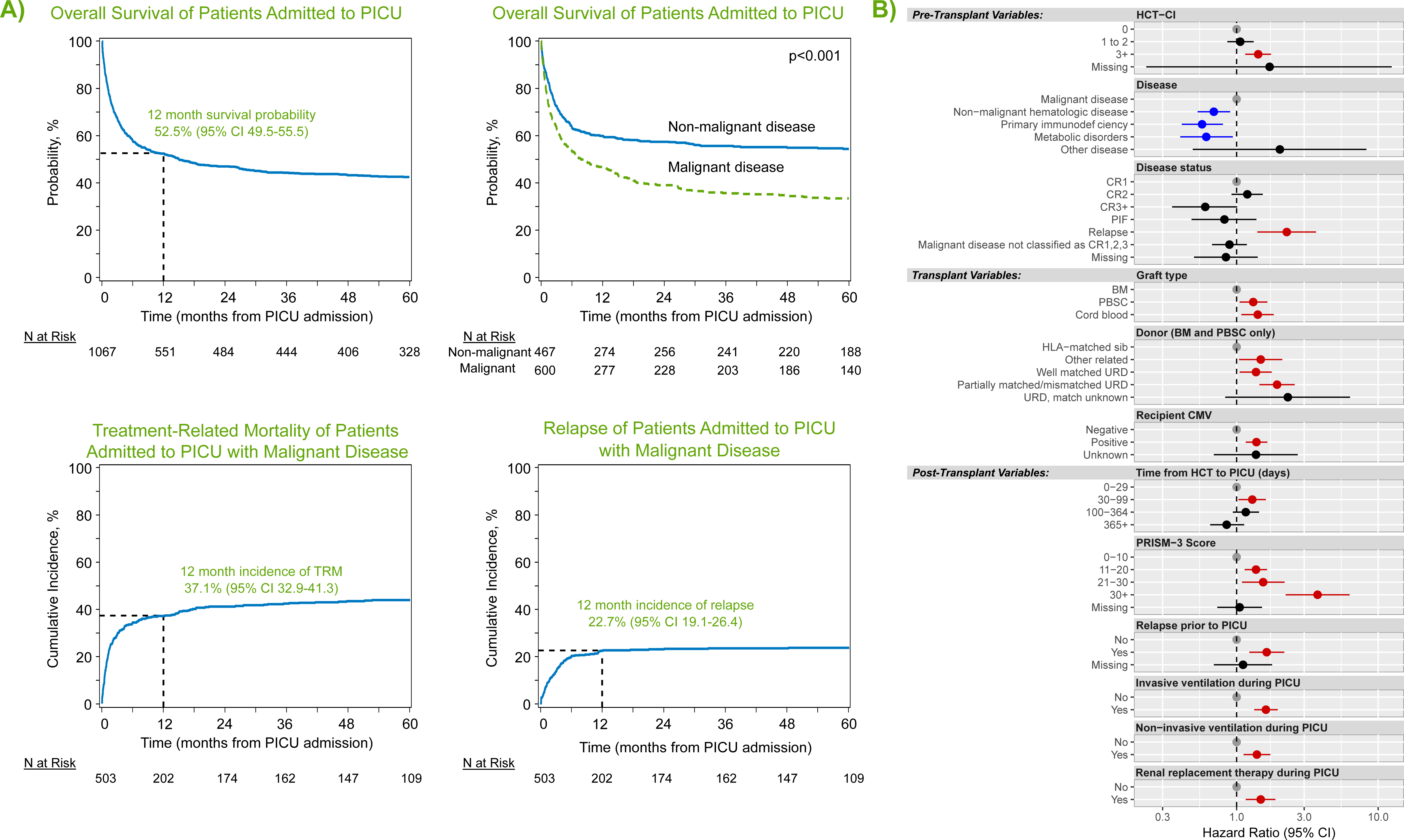
(A) Kaplan Meier estimates of overall survival from the time of PICU admission for all patients (top left) and malignant vs. non-malignant patients (top middle). Cumulative incidence of treatment-related mortality (bottom left) and relapse (bottom middle) among patients transplanted for malignancy are also shown. (B) Factors independently associated with long-term survival from the time of PICU admission to last follow-up in multivariable Cox regression.

On multivariable analysis, we identified 12 risk-factors independently associated with OS among those requiring intensive care (**Figure 2b, Supplemental Table 5)**. Whereas several demographic factors such as age, race, and ethnicity were associated with the need for intensive care admission, once patients were admitted to the ICU, demographics and center size were not independently associated with post-ICU survival. Whereas HCT for inborn errors of metabolism was associated with greater rates of ICU admission, among those admitted to the ICU, HCT for malignant disease was instead associated with worse long-term survival. This was particularly true of malignancy patients with relapsed disease at the time of transplant (HR 2.26, 95% CI 1.40-3.64, p<0.001) or relapsed disease after HCT (HR 1.63, 95% CI 1.23-2.17, p<0.001). In addition to being risk-factors for ICU admission, pre-transplant organ toxicity (HCT-CI ≥3 HR 1.42, 95% CI 1.15-1.75, p<0.001), use of UCB allografts (HR 1.41, 95% CI 1.08-1.83, p=0.012), and allograft donors other than HLA-identical siblings (eg: partially matched unrelated donor HR 1.93, 95% CI 1.45-2.57, p<0.001) were each associated with worse post-ICU survival. Interestingly, although 29% of patients did not have neutrophil engraftment at the time of PICU admission, most of these patients achieved neutrophil engraftment during or after ICU stay, and neutrophil engraftment status at the time of PICU admission was not associated with post-ICU survival (p>0.05). We considered that post-HCT complications may vary according to time post-HCT, with some complications appearing in the early neutropenic phase and others months later. In this analysis, we found that patients who required PICU admission after day +30 had worse long-term survival compared to those who required PICU in the first 30 days (i.e.: PICU admission 30-99 days post-HCT, HR 1.29, 95% CI 1.0-31.61, p=0.029).

### Analysis of 1-year Survivors

Among the subset of the cohort who were alive with follow-up at day +365 (n=5,353 patients), approximately 9% had required intensive care prior to day +365 (n=481). When these 1-year survivors were followed to 5 years post-HCT, the overall survival was approximately 10% lower among those who had required intensive care in the first year (77.1%, 95% CI 73.2-80.8% vs. 87.0%, 95% CI 86.1-88%, p<0.001), corresponding to a hazard ratio of 1.82 (95% CI 1.50-2.22, p<0.001, **Figure 3a, Supplemental Table 6**). Cause-specific mortality among the subset with malignancy showed an approximately 4-fold excess burden of TRM by transplant year 5 in the PICU-exposed group relative to the PICU-unexposed (19.4%, 95% CI 14.1-25.2 vs. 5.6%, 95% CI 4.7-6.6, p<0.001, **Figure 3b**). In contrast, rates of relapse after 1 year were comparable regardless of PICU exposure (**Supplemental Tables 7-8**).

**Figure 3:**
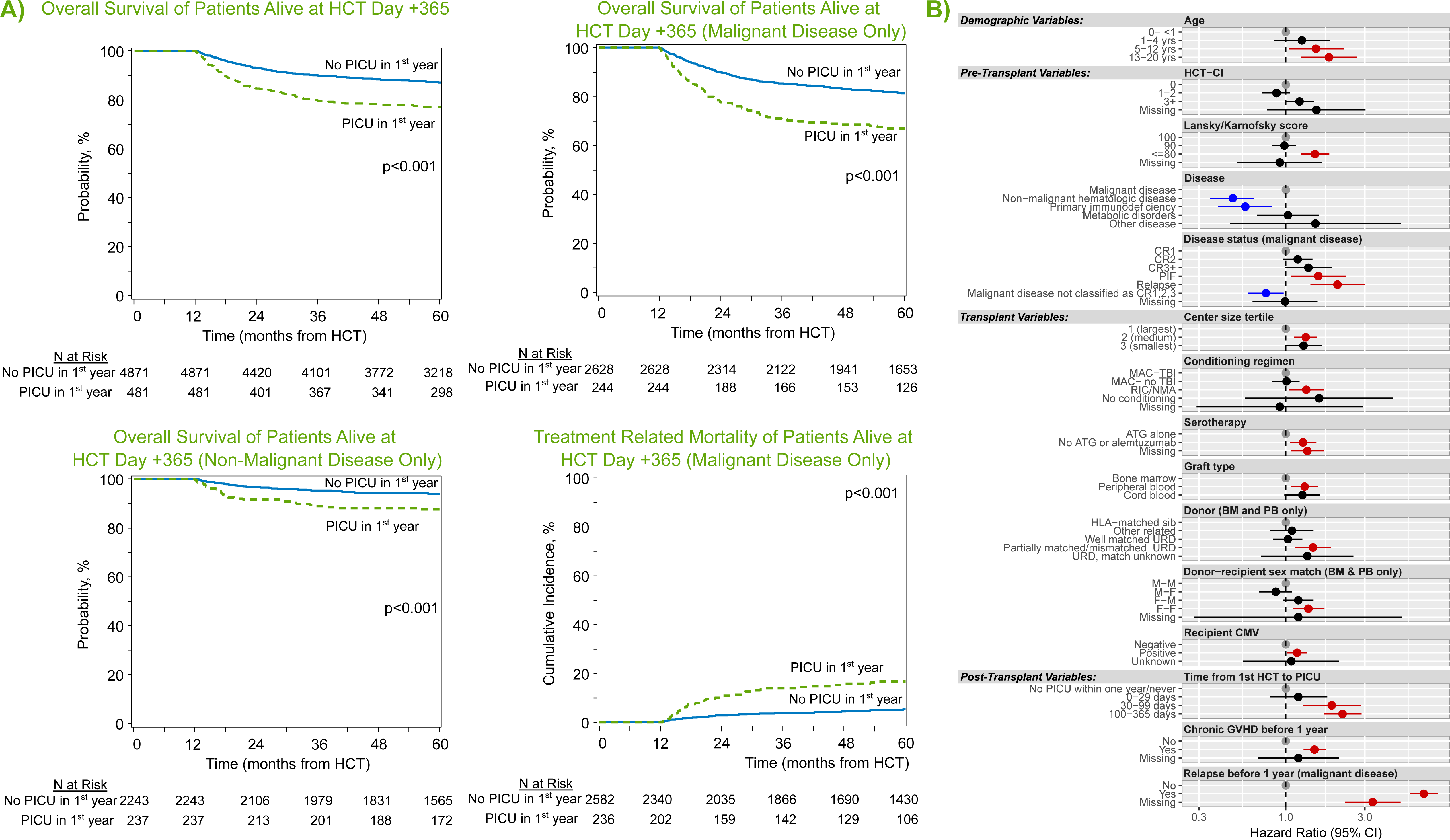
(A) Landmark analysis of only patients surviving to HCT day +365. Kaplan Meier estimates of overall survival from transplant day +365 for all patients (top left), those with malignant disease (top middle), and those with non-malignant disease (bottom left), stratified by need for intensive care in the first year post-HCT. Cumulative incidence of treatment-related mortality (bottom middle) among patients transplanted for malignancy is also shown. (B) Among those alive at HCT day +365, factors independently associated with long-term survival from transplant day +365 to last follow-up in multivariable Cox regression.

On multivariable analysis, the excess mortality after day +365 among PICU-exposed was most pronounced in those who required PICU after day +30. Specifically, PICU admission between HCT day 30-99 (HR 1.89, 95% CI 1.27-2.82) or between HCT day 100-365 (HR 2.20, 95% CI 1.69-2.86, p<0.001) were associated with worse outcomes among 1-year survivors, whereas PICU admission in the first 30 days post-HCT did not have excess mortality beyond non-PICU patients if alive at 1 year (HR 1.19, 95% CI 0.80-1.78, p=0.395). This finding was robust to adjustment for several other factors, including pre-transplant illness severity (HCT-CI, Karnofsky score), transplant variables (recipient CMV+, malignant disease, use of PB or UCB allograft, allograft donor other than HLA-matched sibling), and post-HCT complications (development of cGVHD or relapse prior to one year; **Figure 4c, Supplemental Table 9**).

Organ toxicity data from CIBMTR were available on approximately half of the patients. Patients who required critical care in the first year post-HCT showed a greater prevalence of renal failure, diabetes, liver toxicity, stroke/seizure, and non-infectious pulmonary dysfunction at one-year post-HCT (p<0.001), demonstrating the burden of illness persisted beyond the immediate PICU admission for some patients (**Supplemental Table 10**). Patients who required critical care in the first year post-HCT also showed significantly greater risk of developing new-onset renal failure after day +365 (increase of 4.9% in PICU- exposed vs. 1.6% in PICU non-exposed, p<0.001; **Table 3**). We did not detect a differential increase in the development of diabetes, liver toxicity, stroke/seizure, or non-infectious pulmonary dysfunction after day +365 according to whether PICU was required in the first year, although we could not account for changes or progression in the severity of existing disease.

## DISCUSSION

We report an approximately 15% cumulative incidence of intensive care admission within 5 years following pediatric HCT, with 85% survival to 1^st^ ICU discharge and 52% survival at 1-year post-1^st^ ICU admission. Importantly, ICU survivors alive at 1-year post-HCT still had worse long-term outcomes including OS, TRM, and new dialysis-dependent renal failure when followed to 5 years post-HCT. Together, these data suggest an ongoing burden of toxicity in pediatric HCT patients that continues to limit long-term survival.

First, our report of approximately 8% cumulative incidence of PICU admission within the first 100 days and 15% by year 5 is less than other single-center reports citing 17-35%, which may be due to increased intensity of supportive care outside of the ICU, our exclusion of perioperative and planned ICU admissions, or other factors.^21–23^ Of 69 transplant centers, the third of centers with the highest HCT volume (n=23 centers) accounted for approximately two-thirds of HCT volume and PICU admissions, demonstrating the regionalization of care of these high risk patients. Patients transplanted at a smaller center had increased adjusted risk of ICU admission. The risk for ICU admission associated with younger age and underlying inherited disorders of metabolism may be related to more challenging fluid and airway management in these patients and could be incorporated into patient counseling.^24–26^ The identification of Black and multi-racial background and lower median ZIP code income as risk-factors for post-HCT PICU admission merits further investigation, although these factors were not associated with survival after PICU admission.^27–30^ Finally, the impact of measures of pre-HCT organ toxicity (higher HCT-CI, history of mechanical ventilation) emphasizes the need to optimize organ function in HCT candidates and modify the approach to HCT to increase safety for medically frail patients. Interestingly, time to neutrophil engraftment was not associated with need for intensive care.

Of the patients who required intensive care, we identified a significant discrepancy between survival to ICU discharge (86%) and survival to 1-year post-ICU (53%). Previous reports have suggested ICU survival as low as 20-40%, precluding long-term analyses due to so few survivors.^23, 31–34^ Our data suggest that while survival to ICU discharge is contemporaneously feasible, nearly half of patients required subsequent ICU readmission during the study interval, which could be due to pre-existing medical frailty or ongoing problems such as alloreactivity or poor immune function. As such, survival with future ICU readmission was the most common outcome (44%), ahead of survival without future ICU admission (42%) or ICU death (14%). Therefore, preservation of organ function during each episode of critical illness is crucial to optimizing survival in future episodes of critical illness.^35^ Among ICU patients, those with malignant disease, particularly if relapsed going into or after HCT, were of particularly high risk for poor long-term survival due to the competing risks of relapse and TRM. The PRISM-3 score, a metric of multi-organ dysfunction measured in the first 12 hours of PICU transfer, was strongly associated with long-term outcomes independent of the need for mechanical ventilation or dialysis. While this reiterates that it is important to limit the extent and severity of organ dysfunction, it also indicates the importance of the timing of organ dysfunction relative to ICU care. Studies of early transfer prior to clinical decompensation have shown promising results in reducing adverse events.^36, 37^ Of note, although 26% of patients had not achieved neutrophil engraftment at the time of PICU transfer, this was not associated with worse mortality, and the majority (73%) went on to achieve neutrophil engraftment, suggesting that aggressive supportive care ought not be withheld from patients purely based on neutrophil engraftment status.^38^

Finally, in a landmark analysis of all HCT patients alive at day +365, we found that those who required intensive care in the first year had 10% greater absolute mortality when followed to 5 years post-HCT compared to patients who did not require intensive care in the first year, which was largely attributable to a 4-fold increased risk of TRM. Studies of long-term outcomes in critically ill children are sparse. *Duncan et al* previously showed worse 1-year outcomes in survivors of intensive care; our study extends these findings to 5-years and emphasizes the chronicity of many post-transplant complications.^17^ This risk appeared highest in patients requiring PICU after day +30, again suggesting that transplant complications such as alloreactivity and poor immune function may bear worse prognosis whereas early toxicities related to neutropenia, engraftment, and early fluid overload may be more manageable.

Given the high overall survival among patients alive at day +365, we analyzed the incidence of specific organ toxicities, including development prior to and after day +365. As expected, patients who used intensive care in the first year and survived to day +365 had significantly greater rates of organ toxicity in the first year. Whereas *Broglie et al* recently reported a 12% incidence of non-infectious pulmonary toxicity at 1-year in all pediatric recipients of HCT, our data suggest a much greater burden of approximately 23% at 1-year among those who survived critical illness.^2^ As pulmonary function test (PFT) abnormalities may persist beyond 1-2 years post-HCT in many children, this highlights the need for special attention to follow-up among patients who required intensive care.^3, 39, 40^ However, after 1-year, there was a relatively small proportion of patients who developed new non-infectious organ toxicities, ranging from 0.6-6%. Interestingly, those who required intensive care in the 1^st^ year had a greater incidence of new-onset dialysis-dependent renal failure after day +365, which is consistent with recent reports and suggests a subset of patients with ongoing progression of AKI/chronic kidney disease pathobiology.^41^ Efforts to monitor, mitigate, and address chronic toxicities of pediatric HCT should remain a priority for the field.

This is the largest reported cohort of critically ill pediatric HCT patients with high-quality follow-up to 5 years post-intensive care. Nonetheless, several limitations warrant discussion. First, transplant and ICU practices may have changed since this cohort was merged; further efforts to streamline multi-database merging so as to deliver more contemporary data are needed. Second, repeat HCT could not be differentiated from donor lymphocyte infusion (DLI) in the CIBMTR database and therefore was not addressed. Third, CIBMTR organ toxicity fields are only reported for research-level participants, include somewhat broad categories, and identify the start point of a complication but do not indicate progression or resolution. Fourth, nearly 1,500 patients were excluded from CIBMTR use due to lack of consent, which might introduce selection bias. Fifth, time from first allogeneic HCT to first PICU admission ranged considerably, and the factors associated with early PICU admission (ie.: within 30 days) may be different than those associated with later PICU admission (i.e.: after day +100).

## CONCLUSION

In summary, we report an approximately 15% cumulative incidence of intensive care admission within 5 years after pediatric HCT, with 85% survival to 1^st^ ICU discharge and 52% survival at 1-year post-1^st^ ICU admission. Major risk factors for critical illness included measures of pre-HCT organ toxicity and predisposition to (or development of) post-HCT alloreactivity or malignancy relapse. ICU survivors alive at 1-year post-HCT had worse long-term outcomes including OS, TRM, and new dialysis-dependent renal failure when followed to 5 years post-HCT. However, accrual of other new organ toxicities was minimal among 1-year survivors. These data can be used for patient prognostication and should be targeted in future investigations focused on improving outcomes following pediatric allogeneic HCT.

## Data Use Statement

CIBMTR supports accessibility of research in accord with the National Institutes of Health (NIH) Data Sharing Policy and the National Cancer Institute (NCI) Cancer Moonshot Public Access and Data Sharing Policy. The CIBMTR only releases deDidentified datasets that comply with all relevant global regulations regarding privacy and confidentiality.

## Acknowledgements

VPS data was provided by Virtual Pediatric Systems, (VPS, LLC). No endorsement or editorial restriction of the interpretation of these data or opinions of the authors has been implied or stated. This manuscript has been reviewed by the VPS Research Committee.

