## Supplementary material for "Critical Illness Risk and Long-Term Outcomes Following Intensive Care in Pediatric Hematopoietic Cell Transplant Recipients": Tables

**Table 1. Baseline characteristics of PICU and non-PICU patients receiving allogeneic HCT between 2008-2014 at a center reporting to VPS**

| **Characteristic** | **PICU (n=1,067)** | **Non-PICU (n=5,928)** | **Total (n=6,995)** |
| --- | --- | --- | --- |
| ***Demographics*** | | | |
| **Age at transplant** |  |  |  |
| 0 - <1 | 131 (12) | 534 (9) | 665 (10) |
| 1 – 4 | 250 (23) | 1348 (23) | 1598 (23) |
| 5 – 12 | 370 (35) | 2135 (36) | 2505 (36) |
| 13 – 21 | 316 (30) | 1911 (32) | 2227 (32) |
| **Sex** |  |  |  |
| Male | 624 (59) | 3480 (59) | 4104 (59) |
| Female | 443 (41) | 2448 (41) | 2891 (41) |
| **Race** **^a^** |  |  |  |
| White | 692 (65) | 4316 (73) | 5008 (72) |
| Black or African American | 177 (17) | 810 (14) | 987 (14) |
| Asian | 53 (5) | 303 (5) | 356 (5) |
| Native Hawaiian or other Pacific Islander | 4 (0) | 13 (0) | 17 (0) |
| American Indian or Alaska Native | 9 (1) | 53 (1) | 62 (1) |
| More than one race | 51 (5) | 128 (2) | 179 (3) |
| **Ethnicity** **^a^** |  |  |  |
| Hispanic or Latino | 234 (22) | 1412 (24) | 1646 (24) |
| Not Hispanic or Latino | 808 (76) | 4309 (73) | 5117 (73) |
| Non-resident of the U.S. | 11 (1) | 138 (2) | 149 (2) |
| **Insurance group** **^b^** |  |  |  |
| Private/military/dual insurance **^c^** | 246 (23) | 1555 (26) | 1801 (26) |
| Public insurance only | 180 (17) | 989 (17) | 1169 (17) |
| Uninsured | 10 (1) | 44 (1) | 54 (1) |
| **Median household income by zip code** **^c^** |  |  |  |
| Median (IQR) | 51,985  (40,733-66,239) | 52,393  (41,536-69,484) | 52,348  (41,323-68,775) |
| **BMI status at HCT ^d^** |  |  |  |
| Underweight | 90 (8) | 493 (8) | 583 (8) |
| Normal | 497 (47) | 2965 (50) | 3462 (49) |
| Overweight (adult only) | 10 (1) | 64 (1) | 74 (1) |
| Obese | 114 (11) | 606 (10) | 720 (10) |
| Missing | 356 (33) | 1800 (31) | 2156 (32) |
| ***Pre-transplant-related*** | | | |
| **HCT-CI ^a^** |  |  |  |
| 0 | 671 (63) | 3967 (67) | 4638 (66) |
| 1 | 152 (14) | 758 (13) | 910 (13) |
| 2 | 61 (6) | 323 (5) | 384 (5) |
| 3+ | 181 (17) | 835 (14) | 1016 (15) |
| **Performance score ^a^** |  |  |  |
| 100 | 554 (52) | 3160 (53) | 3714 (53) |
| 90 | 332 (31) | 1832 (31) | 2164 (31) |
| <=80 | 164 (15) | 818 (14) | 982 (14) |
| **History of mechanical ventilation ^a^** |  |  |  |
| No | 918 (86) | 5358 (90) | 6276 (90) |
| Yes | 143 (13) | 511 (9) | 654 (9) |
| **History of invasive fungal infection ^a^** |  |  |  |
| No | 982 (92) | 5472 (92) | 6454 (92) |
| Yes | 81 (8) | 399 (7) | 480 (7) |
| **Disease group** (Subdisease breakdown in Supplemental Table 1) |  |  |  |
| Malignant disease | 600 (56) | 3413 (58) | 4013 (57) |
| Non-malignant hematologic disease | 264 (25) | 1569 (26) | 1833 (26) |
| Primary immunodeficiency | 134 (13) | 671 (11) | 805 (12) |
| Inherited disorders of metabolism | 66 (6) | 258 (4) | 324 (5) |
| Other disease | 3 (0) | 17 (0) | 20 (0) |
| **Malignancy status prior to transplant ^e^** |  |  |  |
| Early disease | 218 (20) | 1280 (22) | 1498 (21) |
| Intermediate disease | 219 (21) | 1264 (21) | 1483 (21) |
| Advanced disease | 52 (5) | 299 (5) | 351 (5) |
| ***Transplant-related*** |  |  |  |
| **Transplant center size** |  |  |  |
| Tertile 1 (>122 allogeneic HCT) | 779 (73) | 3876 (65) | 4655 (67) |
| Tertile 2 (48-122 allogeneic HCT) | 184 (17) | 1626 (27) | 1810 (26) |
| Tertile 3 (< 48 allogeneic HCT) | 104 (10) | 426 (7) | 530 (8) |
| **Time from diagnosis to transplant for malignant diseases only** (months) - median (min-max) | 7 (0-162) | 8 (0-200) | 8 (0-200) |
| **Prescribed conditioning intensity** |  |  |  |
| MAC | 761 (71) | 4367 (74) | 5128 (73) |
| RIC/NMA | 295 (28) | 1498 (25) | 1793 (26) |
| No conditioning | 9 (1) | 48 (1) | 57 (1) |
| **Conditioning intensity/TBI use** |  |  |  |
| MAC-TBI | 345 (32) | 2091 (35) | 2436 (35) |
| MAC-No TBI | 413 (39) | 2262 (38) | 2675 (38) |
| RIC/NMA | 295 (28) | 1498 (25) | 1793 (26) |
| No conditioning | 9 (1) | 48 (1) | 57 (1) |
| **T-cell depletion, CD34+ selection, or ex-vivo expansion** |  |  |  |
| No | 993 (93) | 5622 (95) | 6615 (95) |
| Yes | 72 (7) | 302 (5) | 374 (5) |
| **GVHD prophylaxis** |  |  |  |
| T-cell depletion | 70 (7) | 279 (5) | 349 (5) |
| Post-CY +- other(s) | 7 (1) | 25 (0) | 32 (0) |
| CNI + MTX | 414 (39) | 2902 (49) | 3316 (48) |
| CNI + MMF | 374 (35) | 1473 (29) | 2117 (30) |
| CNI +/- others | 187 (17) | 862 (15) | 1049 (15) |
| Other/unknown | 11 (1) | 98 (2) | 109 (2) |
| No GVHD prophylaxis | 4 (0) | 19 (0) | 23 (0) |
| **ATG/alemtuzumab use** |  |  |  |
| ATG alone | 399 (37) | 2307 (39) | 2706 (39) |
| Alemtuzumab alone | 2 (0) | 10 (0) | 12 (0) |
| No ATG or alemtuzumab | 379 (36) | 2391 (40) | 2770 (40) |
| Missing | 287 (27) | 1220 (21) | 1507 (22) |
| **Graft type** |  |  |  |
| Bone marrow | 601 (56) | 3421 (58) | 4022 (57) |
| Peripheral blood | 155 (15) | 907 (15) | 1062 (15) |
| Umbilical cord blood | 311 (29) | 1600 (27) | 1911 (27) |
| **Donor type (BM and PB only) ^f^** |  |  |  |
| HLA-identical sibling | 186 (17) | 1674 (28) | 1860 (27) |
| Other related | 89 (8) | 340 (6) | 429 (6) |
| Well-matched unrelated (8/8) | 310 (29) | 1578 (27) | 1888 (27) |
| Partially-matched unrelated (7/8)/Mis-matched unrelated (<= 6/8) | 164 (15) | 666 (11) | 830 (12) |
| Unrelated, match unknown | 7 (1) | 66 (1) | 73 (1) |
| **Donor/recipient CMV serostatus ^a^** |  |  |  |
| +/+ | 254 (24) | 1217 (21) | 1471 (21) |
| +/- | 77 (7) | 552 (9) | 629 (9) |
| -/+ | 224 (21) | 1139 (19) | 1363 (19) |
| -/- | 190 (18) | 1346 (23) | 1536 (22) |
| CB - recipient + | 197 (18) | 841 (14) | 1038 (15) |
| CB - recipient - | 109 (10) | 733 (12) | 842 (12) |
| CB - recipient CMV unknown | 5 (0) | 26 (0) | 31 (0) |
| **Donor/recipient ABO match ^b^** |  |  |  |
| Matched | 103 (10) | 729 (12) | 832 (12) |
| Minor mismatch | 46 (4) | 277 (5) | 323 (5) |
| Major mismatch | 37 (3) | 238 (4) | 275 (4) |
| Bi-directional | 11 (1) | 74 (1) | 85 (1) |
| CB | 252 (24) | 1306 (23) | 1558 (23) |
| **Donor/recipient sex match ^a^** |  |  |  |
| M-M | 250 (23) | 1539 (26) | 1789 (26) |
| M-F | 165 (15) | 962 (16) | 1127 (16) |
| F-M | 177 (17) | 1007 (17) | 1184 (17) |
| F-F | 163 (15) | 810 (14) | 973 (14) |
| CB - recipient M | 196 (18) | 927 (16) | 1123 (16) |
| CB - recipient F | 115 (11) | 673 (11) | 788 (11) |

**Legend: Baseline characteristics for all study participants.**

**^a^** Variables with minimal missing data are noted as follows. Race n=386 (6%); Ethnicity n=83 (1%), HCT-CI n=47 (0%); Performance score n=135 (2%); History of mechanical ventilation n=65 (1%); History of proven invasive fungal infection n=61 (1%); Prescribed conditioning intensity n=17 (0%); Conditioning intensity/TBI use n=34 (0%); T-cell depletion n=6 (0%); Donor type n=1 (0%); Donor/recipient CMV serostatus n=85 (1%); Donor/recipient sex match n=11 (0%).

**^b^** Variables with significant missing data are noted as follows. Insurance n=3,971 (57%); Zip code median income n=2,630 (38%); Donor/recipient ABO match n=3922 (56%).

**^c^** There are 8 cases who had both Medicare and Medicaid, these were grouped into dual insurance.

**^d^** Children (age from 2 to <20), BMI is adjusted by sex and age (months) according to CDC smoothed growth charts with percentile cutoffs as follows: underweight (<5%), normal weight (5-84.9%), overweight (85-94.9%), obese (≥95%). Adults 20 years old and older, BMI is interpreted using standard weight status categories as follows: underweight (<18.5), normal weight (18.5-24.9), overweight (25.0-29.9), obese (≥30.0). These categories are the same for men and women of all body types and ages.

**^e^** Does not include NHL/HL, CML, Unknown/MDS.

**^f^** Donor type: Multi-donor n=3 (0%).

**Table 2. Additional Characteristics at PICU Admission (n=1,067)**

| **Characteristic** | **N (%)** |
| --- | --- |
| ***ICU-related*** | |
| **PICU size**, groups created based on total number of admissions |  |
| Tertile 1 (>14 HCT PICU admissions) | 821 (77) |
| Tertile 2 (5-14 HCT PICU admissions) | 175 (17) |
| Tertile 3 (<5 HCT PICU admissions) | 72 (7) |
| **Number of ICU admissions per patient** |  |
| 1 | 597 (56) |
| 2 | 248 (23) |
| 3+ | 222 (21) |
| **Age at 1st PICU admission (years) ^a^** |  |
| 0 - <1 | 98 (9) |
| 1 – 4 | 259 (24) |
| 5 – 12 | 365 (34) |
| 13 – 20 | 344 (32) |
| **BMI status at PICU admission ^c^** |  |
| Underweight | 90 (8) |
| Normal | 497 (47) |
| Overweight (adult only) | 10 (1) |
| Obese | 114 (11) |
| Missing | 356 (33) |
| **PRISM-3 Score (median, IQR)** | 9 (4-14) |
| **Critical Care Therapies in 1^st^ ICU admission** |  |
| Invasive mechanical ventilation | 395 (37) |
| Noninvasive mechanical ventilation | 175 (16) |
| Renal replacement therapy | 126 (12) |
| **Infections in 1^st^ ICU admission** |  |
| Gram positive infection | 115 (11) |
| Gram negative infection | 87 (8) |
| Fungal infection | 94 (9) |
| Viral infection | 273 (25) |
| ***Transplant-Related*** | |
| **Time from HCT to 1st ICU admission** (months) - median (min-max) | 3 (0-70) |
| **Neutrophil recovery relationship to 1st ICU admission ^a^** |  |
| Neutrophil engrafted prior to 1st ICU admission | 761 (71) |
| Neutrophil engrafted after 1st ICU admission and before 1st ICU discharge | 89 (8) |
| Neutrophil engrafted after ICU discharge | 122 (11) |
| Never achieved neutrophil engraftment | 77 (7) |
| **aGVHD relationship to 1st ICU admission ^a,b^** |  |
| No aGVHD | 299 (65) |
| aGVHD prior to 1st ICU admission | 124 (27) |
| Grade I-II | 53 (11) |
| Grade III-IV | 69 (15) |
| aGVHD after 1st ICU admission and before 1st ICU discharge | 12 (3) |
| Grade I-II | 3 (0) |
| Grade III-IV | 9 (2) |
| aGVHD after 1st ICU discharge | 27 (6) |
| Grade I-II | 11 (2) |
| Grade III-IV | 16 (4) |
| **cGVHD relationship to 1st ICU admission ^a^** |  |
| No CGVHD | 748 (70) |
| CGVHD prior to 1st ICU admission | 130 (12) |
| Limited | 33 (3) |
| Extensive | 94 (9) |
| CGVHD post 1st ICU admission | 163 (15) |
| Limited | 43 (4) |
| Extensive | 119 (11) |
| **Malignancy Relapse prior to 1st ICU visit** | 103 (10) |

**Legend: Additional characteristics at time of PICU admission.**

**^a^** Variables with minimal missing data are noted as follows. Age at 1^st^ PICU admission n=1 (0%); Neutrophil recovery relationship to 1^st^ PICU admission n=18 (2%); aGVHD grade unknown n=2 (0%); cGVHD relationship to 1^st^ PICU admission: Missing n=26 (2%), cGVHD grade unknown n=4 (0%).

**^b^** Variables with significant missing data are noted as follows. Insurance n=3,971 (57%); Zip code median income n=2,630 (38%); Donor/recipient ABO match n=3922 (56%); aGVHD relationship to 1^st^ PICU admission: Missing or not collected n=605 (56%).

^c^ Children (age from 2 to <20), BMI is adjusted by sex and age (months) according to CDC smoothed growth charts with percentile cutoffs as follows: underweight (<5%), normal weight (5-84.9%), overweight (85-94.9%), obese (≥95%). Adults 20 years old and older, BMI is interpreted using standard weight status categories as follows: underweight (<18.5), normal weight (18.5-24.9), overweight (25.0-29.9), obese (≥30.0). These categories are the same for men and women of all body types and ages.

**Table 3. Reported organ toxicities among 1-year HCT survivors**

|  | **Alive at Day +365 without PICU (N = 2157)** | | **Alive at Day +365 with PICU**  **(N = 214)** | |  |
| --- | --- | --- | --- | --- | --- |
| **Outcomes** | **N** | **Prob (95% CI)** | **N** | **Prob (95% CI)** | **P Value** |
| **Congestive heart failure** | 2116 |  | 206 |  |  |
| 1-year | 2106 | 0.5 (0.3-0.9)% | 206 | 0.5 (0-1.9)% |  |
| 2-year | 1912 | 0.8 (0.5-1.2)% | 171 | 1 (0.1-2.8)% | 0.769 |
| 3-year | 1789 | 1 (0.6-1.5)% | 155 | 1 (0.1-2.8)% |  |
| 5-year | 1435 | 1.2 (0.7-1.7)% | 126 | 1 (0.1-2.8)% |  |
| **Renal failure requiring dialysis** | 2113 |  | 206 |  |  |
| 1-year | 2081 | 1.6 (1.1-2.1)% | 187 | 9.7 (6-14.1)% |  |
| 2-year | 1889 | 2.7 (2-3.4)% | 153 | 13.6 (9.3-18.6)% | <0.001 |
| 3-year | 1768 | 2.9 (2.2-3.6)% | 138 | 14.6 (10.1-19.8)% |  |
| 5-year | 1420 | 3.2 (2.5-4)% | 111 | 14.6 (10.1-19.8)% |  |
| **Diabetes/hyperglycemia** | 2026 |  | 200 |  |  |
| 1-year | 1923 | 5.1 (4.2-6.1)% | 174 | 13.5 (9.1-18.6)% |  |
| 2-year | 1738 | 5.9 (4.9-6.9)% | 142 | 14 (9.5-19.2)% | 0.371 |
| 3-year | 1622 | 6 (5-7.1)% | 127 | 14 (9.5-19.2)% |  |
| 5-year | 1309 | 6.5 (5.4-7.6)% | 105 | 14 (9.5-19.2)% |  |
| **Non-infectious liver toxicity** | 2112 |  | 205 |  |  |
| 1-year | 1718 | 18.7 (17.1-20.4)% | 146 | 29.3 (23.2-35.7)% |  |
| 2-year | 1543 | 20.7 (19-22.4)% | 118 | 30.7 (24.6-37.2)% | 0.683 |
| 3-year | 1435 | 21.2 (19.5-23)% | 107 | 31.3 (25.1-37.8)% |  |
| 5-year | 1102 | 22 (20.3-23.8)% | 82 | 31.3 (25.1-37.8)% |  |
| **Stroke/seizure** | 2116 |  | 205 |  |  |
| 1-year | 2051 | 3.1 (2.4-3.9)% | 175 | 15.1 (10.5-20.4)% |  |
| 2-year | 1846 | 4.6 (3.7-5.5)% | 146 | 17.1 (12.2-22.5)% | 0.081 |
| 3-year | 1723 | 5.1 (4.2-6.1)% | 132 | 18.6 (13.6-24.2)% |  |
| 5-year | 1336 | 5.5 (4.6-6.6)% | 105 | 18.6 (13.6-24.2)% |  |
| **Non-infectious pulmonary dysfunction** | 1987 |  | 175 |  |  |
| 1-year | 1845 | 7.2 (6.1-8.4)% | 136 | 22.9 (16.9-29.4)% |  |
| 2-year | 1656 | 9.9 (8.6-11.2)% | 109 | 27.5 (21.1-34.4)% | 0.270 |
| 3-year | 1550 | 10.4 (9.1-11.7)% | 102 | 27.5 (21.1-34.4)% |  |
| 5-year | 1252 | 10.7 (9.4-12.1)% | 87 | 27.5 (21.1-34.4)% |  |

**Legend: Organ toxicities reported in CIBMTR (evaluable for the data-intensive CRF track only).** P-values are based on significance tests comparing the cumulative incidence between the two sub-populations through years 2-5. Incidence of late effects within the first year does not have an impact on this p-value. 1-year estimates are reported as descriptive estimates only.
