## Supplement for "Critical Illness Risk and Long-Term Outcomes Following Intensive Care in Pediatric Hematopoietic Cell Transplant Recipients"

**Contents:**

**Supplemental Table 1.** Baseline disease of PICU and non-PICU patients receiving alloHCT between 2008-2014 at a center reporting to VPS

**Supplemental Table 2.** Multivariable model for PICU admission (N=6991)

**Supplemental Table 3.** Univariate Analysis of Outcomes (Only patients admitted to PICU, all indications)

**Supplemental Table 4.** Univariate Analysis of Outcomes (Only patients admitted to PICU, malignant patients only)

**Supplemental Table 5.** Multivariable model for Survival of patients with PICU admission (N=1067)

**Supplemental Table 6.** Univariate Analysis of Outcomes (Only those who survived to post HCT Day 365+)

**Supplemental Table 7.** Univariate Analysis of Outcomes (Only those who survived to post HCT day 365+, malignant patients only)

**Supplemental Table 8.** Univariate Analysis of Outcomes (Only those who survived without relapse to post HCT Day 365+, malignant patients only)

**Supplemental Table 9.** Multivariate model for Survival of patients who survived to post-HCT day +365 (N=5340)

**Supplemental Table 10.** Late Effects (CRF subset)

**Supplemental Table 1. Baseline disease of PICU and non-PICU patients receiving alloHCT between 2008-2014 at a center reporting to VPS**

| **Disease - no. (%)** | **PICU**  **(n=1,067)** | **Non-PICU**  **(n=5,928)** | **Total**  **(n=6,995)** |
| --- | --- | --- | --- |
| Malignant disease |  |  |  |
| AML | 203 (19) | 1172 (20) | 1375 (20) |
| ALL | 237 (22) | 1444 (24) | 1681 (24) |
| Other leukemia ^a^ | 0 (0) | 2 (0) | 2 (0) |
| CML | 20 (2) | 95 (2) | 115 (2) |
| MDS/MPD | 76 (7) | 364 (6) | 440 (6) |
| Other acute leukemia ^b^ | 26 (2) | 114 (2) | 140 (2) |
| NHL | 21 (2) | 155 (3) | 176 (3) |
| HL | 13 (1) | 52 (1) | 65 (1) |
| PCD/MM | 0 (0) | 2 (0) | 2 (0) |
| Other Malignancies | 4 (0) | 13 (0) | 17 (0) |
| Non-malignant hematologic disease |  |  |  |
| SAA | 60 (6) | 565 (10) | 625 (9) |
| Inherited abnormalities erythrocyte differentiation or function | 123 (11) | 718 (12) | 841 (12) |
| Inherited abnormalities of platelets | 0 (0) | 44 (1) | 44 (1) |
| Histiocytic disorders | 81 (8) | 242 (4) | 323 (5) |
| Primary immunodeficiency |  |  |  |
| SCID and other immune system disorders | 128 (12) | 655 (11) | 783 (11) |
| Autoimmune Diseases | 6 (1) | 16 (0) | 22 (0) |
| Metabolic disorders |  |  |  |
| Inherited disorders of metabolism | 66 (6) | 258 (4) | 324 (5) |
| Other disease |  |  |  |
| Other, specify ^c^ | 3 (0) | 17 (0) | 20 (0) |

**Legend:** Detailed underlying diagnoses of study participants as listed in CIBMTR.

**^a^** Other leukemia, specify: large granulomatous lyphocytosis disorder (n=1), NK cell (n=1).

**^b^** Other acute leukemia, specify: Acute undifferentiated leukemia (n=17), biphenotypic, binleaneage or hybrid leukemia (n=109), blastic plasmacytoid dendritic cell neoplasm (n=1), acute leukemia of ambiguous lineage (n=1), APML (n=1), blastic plasmacytoid dendritic cell leukemia (n=1), blastic plasmacytoid dendritic cell neoplasm (n=2) mixed phenotype (n=1), NK AML (n=1), NK cell leukemia (n=2), NK leukemia (n=1), PH+ ALL (n=1), plasmocytoid dendritic cell neoplasm (n=1), tri-linage leukemia (n=1).

**^c^** Other specify: Epidermolysis bullosa (n=11), CEP (n=1), Chronic EBV secondary to EBV associated Hodgkin lymphoma (n=1), Congenital neutropenia (n=1), Dyskeratosis congenita (n=1), Erythropoietic protoporphyria (n=1), Hoyeraal_Hreidarsson syndrome/dyskeratosis congenita (n=1), Infantile neuroaxonal dysyroph (n=1), Nijemann breakage syndrome (n=1), NK cell SCAEBV (n=1) .

**Supplemental Table 2. Multivariable model for PICU admission (N=6991)**

| **Parameter** | **Category** | **Freq** | **HR** | **CI lower** | **CI upper** | **p-value** |
| --- | --- | --- | --- | --- | --- | --- |
| ***Demographics*** | | | | | | |
| Age | 0-0.99 | 665 | 1.00 | . | . | 0.0337 |
|  | 1-3.99 | 1598 | 0.77 | 0.61 | 0.96 | 0.0228 |
|  | 5-12.99 | 2504 | 0.73 | 0.58 | 0.91 | 0.0062 |
|  | 13-20.99 | 2224 | 0.70 | 0.55 | 0.90 | 0.0047 |
| Race | White | 4941 | 1.00 | . | . | <.0001 |
|  | Black | 983 | 1.33 | 1.11 | 1.59 | 0.0019 |
|  | Asian | 341 | 1.08 | 0.81 | 1.44 | 0.6132 |
|  | Other | 76 | 1.24 | 0.71 | 2.16 | 0.4447 |
|  | More than one race | 177 | 2.00 | 1.50 | 2.67 | <.0001 |
|  | Non-US resident | 149 | 0.62 | 0.34 | 1.15 | 0.1303 |
|  | Missing | 324 | 1.95 | 1.51 | 2.51 | <.0001 |
| Ethnicity (US only) | Non-Hispanic or Latino | 5114 | 1.00 | . | . | 0.1427 |
|  | Hispanic or Latino | 1645 | 0.85 | 0.72 | 1.00 | 0.0492 |
|  | Missing | 83 | 0.89 | 0.51 | 1.54 | 0.6698 |
| Median Household Income by ZIP code | <$75,000 | 3559 | 1.00 | . | . | <.0001 |
|  | >= $75,000 | 809 | 0.66 | 0.53 | 0.83 | 0.0004 |
|  | Missing | 2623 | 1.13 | 0.98 | 1.29 | 0.0879 |
| ***Pre-transplant*** | | | | | | |
| HCT-CI | 0 | 4637 | 1.00 | . | . | 0.0359 |
|  | 1-2 | 1292 | 1.09 | 0.93 | 1.28 | 0.2816 |
|  | 3+ | 1016 | 1.26 | 1.06 | 1.49 | 0.0084 |
|  | Missing | 46 | 0.41 | 0.09 | 1.89 | 0.2555 |
| History of mechanical ventilation | No | 6272 | 1.00 | . | . | 0.0001 |
|  | Yes | 654 | 1.49 | 1.24 | 1.79 | <.0001 |
|  | Unknown | 65 | 0.95 | 0.40 | 2.30 | 0.9181 |
| Disease | Malignant disease | 4010 | 1.00 | . | . | 0.0040 |
|  | Non-malignant hematologic | 1833 | 1.02 | 0.87 | 1.20 | 0.8156 |
|  | Primary immunodeficiency | 804 | 0.97 | 0.77 | 1.21 | 0.7600 |
|  | Metabolic disorders | 324 | 1.69 | 1.28 | 2.23 | 0.0002 |
|  | Other disease | 20 | 1.16 | 0.37 | 3.63 | 0.8007 |
| ***Transplant related*** | | | | | | |
| Center size tertile | 1 (largest) | 4653 | 1.00 | . | . | <.0001 |
|  | 2 | 1809 | 0.64 | 0.54 | 0.75 | <.0001 |
|  | 3 (smallest) | 529 | 1.32 | 1.07 | 1.63 | 0.0092 |
| GVHD prophylaxis | CNI + MTX | 3313 | 1.00 | . | . | <.0001 |
|  | TCD | 348 | 1.62 | 1.17 | 2.23 | 0.0034 |
|  | Post Cy +/- other | 32 | 1.31 | 0.60 | 2.85 | 0.5032 |
|  | CNI + MMF | 2117 | 1.40 | 1.19 | 1.65 | <.0001 |
|  | CNI +/- other | 1049 | 1.50 | 1.25 | 1.81 | <.0001 |
|  | No GVHD prophylaxis | 23 | 1.81 | 0.67 | 4.89 | 0.2451 |
|  | Other/unknown | 109 | 0.93 | 0.51 | 1.70 | 0.8072 |
| Graft type | Bone marrow | 4020 | 1.00 | . | . | <.0001 |
|  | Peripheral blood | 1060 | 0.76 | 0.61 | 0.94 | 0.0114 |
|  | Cord blood | 1911 | 1.46 | 1.18 | 1.81 | 0.0005 |
| Donor (BM and PB only) | HLA-identical sib | 1860 | 1.00 | . | . | <.0001 |
|  | Other related | 429 | 2.00 | 1.51 | 2.67 | <.0001 |
|  | Well matched URD | 1888 | 1.75 | 1.45 | 2.12 | <.0001 |
|  | Partially matched URD | 830 | 1.95 | 1.57 | 2.43 | <.0001 |
|  | URD, match unknown | 73 | 0.95 | 0.44 | 2.04 | 0.8977 |
| Recipient CMV | Negative | 3006 | 1.00 | . | . | <.0001 |
|  | Positive | 3869 | 1.46 | 1.28 | 1.67 | <.0001 |
|  | Unknown | 116 | 0.90 | 0.53 | 1.51 | 0.6807 |
| ***Post-transplant*** | | | | | | |
| Acute GVHD 3-4 | No | 5745 | 1.00 | . | . | 0.0001 |
|  | Yes | 474 | 1.65 | 1.28 | 2.14 | 0.0001 |
|  | Missing | 568 | 1.69 | 1.43 | 2.01 | <.0001 |
| Chronic GVHD | None/Limited | 5791 | 1.00 | . | . | <.0001 |
|  | Extensive | 1134 | 1.80 | 1.43 | 2.28 | <.0001 |
|  | Missing | 66 | 1.14 | 0.75 | 1.74 | 0.5271 |
| Relapse (malignant diseases only) | No | 2758 | 1.00 | . | . | <.0001 |
|  | Yes | 1133 | 6.26 | 5.00 | 7.84 | <.0001 |
|  | Missing | 119 | 1.47 | 0.96 | 2.24 | 0.0779 |

**Legend:** Multivariable competing risk regression for the outcome of PICU admission with death without PICU admission as a competing risk.

**Supplemental Table 3. Univariate Analysis of Outcomes (Only patients admitted to PICU, all indications)**

| **Outcomes** | **N** | **Prob (95% CI)** |
| --- | --- | --- |
| Overall survival | 1067 |  |
| 1-year |  | 52.5 (49.5-55.5)% |
| 3-year |  | 44.4 (41.5-47.5)% |
| 5-year |  | 42.6 (39.6-45.6)% |

**Legend:** Survival time from 1^st^ PICU admission.

**Supplemental Table 4. Univariate Analysis of Outcomes (Only patients admitted to PICU, malignant patients only)**

| **Outcomes** | **N** | **Prob (95% CI)** |
| --- | --- | --- |
| Overall survival | 601 |  |
| 1-year |  | 46.7 (42.7-50.7)% |
| 3-year |  | 35.8 (32-39.7)% |
| 5-year |  | 33.5 (29.8-37.4)% |
| Treatment related mortality | 508 |  |
| 1-year |  | 37.1 (32.9-41.3)% |
| 3-year |  | 42.3 (38-46.6)% |
| 5-year |  | 43.9 (39.5-48.2)% |
| Relapse/Progression | 508 |  |
| 1-year |  | 22.7 (19.1-26.4)% |
| 3-year |  | 23.7 (20.1-27.5)% |
| 5-year |  | 23.9 (20.3-27.7)% |
| Disease free survival | 508 |  |
| 1-year |  | 40.2 (36-44.6)% |
| 3-year |  | 34 (29.9-38.2)% |
| 5-year |  | 32.2 (28.2-36.4)% |

**Legend:** Outcomes from the time of 1^st^ PICU admission, for malignancy patients only.

**Supplemental Table 5. Multivariable model for Survival of patients with PICU admission, N=1067**

| **Parameter** | **Category** | **Freq** | **HR** | **CI lower** | **CI upper** | **p-value** |
| --- | --- | --- | --- | --- | --- | --- |
| ***Pre-transplant*** | | | | | | |
| HCT-CI | 0 | 671 | 1.00 | . | . | 0.0102 |
|  | 1-2 | 213 | 1.06 | 0.86 | 1.32 | 0.5754 |
|  | 3+ | 181 | 1.42 | 1.15 | 1.75 | 0.0009 |
|  | Missing | 2 | 1.71 | 0.23 | 12.46 | 0.5960 |
| Disease | Malignant disease | 600 | 1.00 | . | . | 0.0026 |
|  | Non-malignant hematologic | 264 | 0.69 | 0.53 | 0.90 | 0.0059 |
|  | Primary immunodeficiency | 134 | 0.57 | 0.41 | 0.80 | 0.0009 |
|  | Metabolic disorders | 66 | 0.61 | 0.40 | 0.94 | 0.0252 |
|  | Other disease | 3 | 2.02 | 0.49 | 8.26 | 0.3300 |
| Disease status | CR1 | 187 | 1.00 | . | . | 0.0011 |
|  | CR2 | 173 | 1.19 | 0.92 | 1.53 | 0.1883 |
|  | CR3+ | 30 | 0.60 | 0.35 | 1.01 | 0.0556 |
|  | PIF | 26 | 0.82 | 0.48 | 1.38 | 0.4489 |
|  | Relapse | 24 | 2.26 | 1.40 | 3.64 | 0.0008 |
|  | Malignant disease not classified as CR1,2,3 | 134 | 0.89 | 0.67 | 1.18 | 0.4001 |
|  | Missing | 26 | 0.84 | 0.50 | 1.41 | 0.5063 |
| ***Transplant related*** | | | | | | |
| Graft type | BM | 601 | 1.00 | . | . | 0.0042 |
|  | PBSC | 155 | 1.31 | 1.05 | 1.65 | 0.0192 |
|  | Cord blood | 311 | 1.41 | 1.08 | 1.83 | 0.0117 |
| Donor (BM and PBSC only) | HLA-matched sib | 186 | 1.00 | . | . | 0.0002 |
|  | Other related | 89 | 1.48 | 1.04 | 2.10 | 0.0280 |
|  | Well matched URD | 310 | 1.37 | 1.05 | 1.77 | 0.0189 |
|  | Partially matched URD | 164 | 1.93 | 1.45 | 2.57 | <.0001 |
|  | URD, match unknown | 7 | 2.30 | 0.83 | 6.33 | 0.1077 |
| Recipient CMV | Negative | 376 | 1.00 | . | . | 0.0016 |
|  | Positive | 675 | 1.38 | 1.16 | 1.65 | 0.0004 |
|  | Unknown | 16 | 1.37 | 0.69 | 2.70 | 0.3691 |
| ***Post-transplant*** | | | | | | |
| Time b/w HCT and PICU, days | 0-29 | 348 | 1.00 | . | . | 0.0157 |
|  | 30-99 | 234 | 1.29 | 1.03 | 1.61 | 0.0291 |
|  | 100-364 | 314 | 1.16 | 0.94 | 1.44 | 0.1677 |
|  | 365+ | 171 | 0.85 | 0.65 | 1.13 | 0.2615 |
| PRISM 3 | 0-10 | 558 | 1.00 | . | . | <.0001 |
|  | 11-20 | 374 | 1.37 | 1.14 | 1.64 | 0.0007 |
|  | 21-30 | 55 | 1.54 | 1.09 | 2.18 | 0.0147 |
|  | 30+ | 18 | 3.73 | 2.22 | 6.28 | <.0001 |
|  | Missing | 62 | 1.05 | 0.73 | 1.51 | 0.7846 |
| Relapse prior to PICU | No | 474 | 1.00 | . | . | 0.0036 |
|  | Yes | 103 | 1.63 | 1.23 | 2.17 | 0.0008 |
|  | Missing | 23 | 1.11 | 0.69 | 1.78 | 0.6571 |
| Invasive vent after admission to PICU | No | 672 | 1.00 | . | . | <.0001 |
|  | Yes | 395 | 1.61 | 1.33 | 1.95 | <.0001 |
| Non-invasive vent after admission to PICU | No | 892 | 1.00 | . | . | 0.0031 |
|  | Yes | 175 | 1.39 | 1.12 | 1.73 | 0.0031 |
| Renal replacement therapy during 1^st^ PICU | No | 941 | 1.00 | . | . | 0.0016 |
|  | Yes | 126 | 1.48 | 1.16 | 1.88 | 0.0016 |

**Legend:** Multivariable model for long-term survival among PICU patients (from the time of PICU admission).

**Supplemental Table 6. Univariate Analysis of Outcomes (Only those who survived to post HCT Day 365+)**

|  | **Alive at 365 without PICU (N = 4872)** | | **Alive at 365 with PICU (N = 481)** | |  |
| --- | --- | --- | --- | --- | --- |
| **Outcomes** | **N** | **Prob (95% CI)** | **N** | **Prob (95% CI)** | **P Value** |
| Overall survival | 4872 |  | 481 |  | <0.001 |
| 1-year |  | 100% |  | 100% |  |
| 3-year |  | 89.9 (89-90.7)% |  | 79.6 (75.9-83.1)% |  |
| 5-year |  | 87.0 (86.1-88)% |  | 77.1 (73.2-80.8)% |  |

**Legend:** Comparison of long-term survival among patients who did vs. did not require PICU in the first year post-HCT.

**Supplemental Table 7. Univariate Analysis of Outcomes (Only those who survived to post HCT day 365+, malignant patients only)**

|  | **Alive at 365 without PICU (N = 2628)** | | **Alive at 365 with PICU (N = 244)** | |  |
| --- | --- | --- | --- | --- | --- |
| **Outcomes** | **N** | **Prob (95% CI)** | **N** | **Prob (95% CI)** | **P Value** |
| Overall survival | 2628 |  | 244 |  | <0.001 |
| 1-year | 2628 | 100% | 244 | 100% |  |
| Treatment related mortality | 2582 |  | 236 |  | <0.001 |
| 1-year | 2341 | 0% | 203 | 0% |  |
| Relapse | 2582 |  | 236 |  | 0.451 |
| 1-year | 2341 | 9.4 (8.3-10.5)% | 203 | 14.4 (10.2-19.2)% |  |
| Disease free survival | 2582 |  | 236 |  | <0.001 |
| 1-year | 2340 | 90.6 (89.5-91.7)% | 202 | 85.6 (80.8-89.8)% |  |

**Legend:** Comparison of long-term outcomes among patients who did vs. did not require PICU in the first year post-HCT (malignancy patients only).

**Supplemental Table 8. Univariate Analysis of Outcomes (Only those who survived without relapse to post HCT Day 365+, malignant patients only)**

|  | **Alive at 365 without PICU (N = 2340)** | | **Alive at 365 with PICU (N = 202)** | |  |
| --- | --- | --- | --- | --- | --- |
| **Outcomes** | **N** | **Prob (95% CI)** | **N** | **Prob (95% CI)** | **P Value** |
| Overall survival | 2340 |  | 202 |  | <0.001 |
| 2-year | 2156 | 94 (93-94.9)% | 167 | 83.6 (78.1-88.4)% |  |
| 3-year | 1990 | 89.8 (88.6-91)% | 150 | 77.4 (71.4-83)% |  |
| 5-year | 1548 | 85.9 (84.4-87.3)% | 114 | 73 (66.6-79)% |  |
| Treatment related mortality | 2340 |  | 202 |  | <0.001 |
| 2-year | 2037 | 3 (2.3-3.7)% | 160 | 12.4 (8.2-17.4)% |  |
| 3-year | 1868 | 4.1 (3.4-5)% | 143 | 16 (11.3-21.5)% |  |
| 5-year | 1430 | 5.6 (4.7-6.6)% | 106 | 19.4 (14.1-25.2)% |  |
| Relapse | 2340 |  | 202 |  | 0.223 |
| 2-year | 2037 | 8.3 (7.2-9.4)% | 160 | 8 (4.6-12.1)% |  |
| 3-year | 1868 | 11.5 (10.2-12.8)% | 143 | 10.5 (6.6-15.1)% |  |
| 5-year | 1430 | 14.5 (13.1-16)% | 106 | 11.6 (7.5-16.4)% |  |
| Disease free survival | 2340 |  | 202 |  | <0.001 |
| 2-year | 2035 | 88.7 (87.4-90)% | 159 | 79.6 (73.8-84.9)% |  |
| 3-year | 1866 | 84.3 (82.8-85.8)% | 142 | 73.5 (67.1-79.3)% |  |
| 5-year | 1430 | 79.7 (78-81.4)% | 106 | 69.1 (62.4-75.3)% |  |

**Legend:** Comparison of long-term outcomes among patients who did vs. did not require PICU in the first year post-HCT (malignancy patients without relapse in the first 365 days only).

**Supplemental Table 9. Multivariate model for Survival of patients who survived to post-HCT day +365 (N=5340)**

| **Parameter** | **Category** | **Freq** | **HR** | **CI lower** | **CI upper** | **p-value** |
| --- | --- | --- | --- | --- | --- | --- |
| ***Demographics*** | | | | | | |
| Age | 0- <1 | 509 | 1.00 | . | . | 0.0011 |
|  | 1-4 yrs | 1205 | 1.25 | 0.85 | 1.84 | 0.2596 |
|  | 5-12 yrs | 1987 | 1.52 | 1.04 | 2.23 | 0.0290 |
|  | 13-20 yrs | 1639 | 1.82 | 1.23 | 2.68 | 0.0026 |
| ***Pre-transplant*** | | | | | | |
| HCT CI | 0 | 3641 | 1.00 | . | . | 0.0400 |
|  | 1-2 | 973 | 0.88 | 0.72 | 1.06 | 0.1857 |
|  | 3+ | 690 | 1.21 | 0.99 | 1.48 | 0.0644 |
|  | Missing | 36 | 1.53 | 0.77 | 3.02 | 0.2211 |
| Lansky/Karnofsky score | 100 | 2947 | 1.00 | . | . | 0.0002 |
|  | 90 | 1635 | 0.98 | 0.83 | 1.15 | 0.7998 |
|  | <=80 | 656 | 1.50 | 1.24 | 1.83 | <.0001 |
|  | Missing | 102 | 0.92 | 0.51 | 1.65 | 0.7776 |
| Disease | Malignant disease | 2870 | 1.00 | . | . | <.0001 |
|  | Non-malignant hematologic | 1557 | 0.48 | 0.35 | 0.64 | <.0001 |
|  | Primary immunodeficiency | 662 | 0.57 | 0.39 | 0.83 | 0.0038 |
|  | Metabolic disorders | 240 | 1.03 | 0.67 | 1.59 | 0.8816 |
|  | Other disease | 11 | 1.51 | 0.46 | 4.94 | 0.4924 |
| Disease status (malignant disease) | CR1 | 999 | 1.00 | . | . | <.0001 |
|  | CR2 | 826 | 1.18 | 0.96 | 1.45 | 0.1157 |
|  | CR3+ | 157 | 1.37 | 0.99 | 1.90 | 0.0554 |
|  | PIF | 87 | 1.57 | 1.07 | 2.31 | 0.0214 |
|  | Relapse | 80 | 2.05 | 1.41 | 3.00 | 0.0002 |
|  | Malignant disease not classified as CR1,2,3 | 608 | 0.76 | 0.59 | 0.97 | 0.0270 |
|  | Missing | 113 | 0.99 | 0.63 | 1.55 | 0.9623 |
| ***Transplant related*** | | | | | | |
| Center size tertile | 1 | 3612 | 1.00 | . | . | 0.0015 |
|  | 2 | 1341 | 1.32 | 1.12 | 1.54 | 0.0007 |
|  | 3 | 387 | 1.28 | 0.99 | 1.65 | 0.0595 |
| Conditioning | MAC-TBI | 1785 | 1.00 | . | . | 0.1175 |
|  | MAC- no TBI | 2034 | 1.01 | 0.83 | 1.21 | 0.9570 |
|  | RIC/NMA | 1450 | 1.33 | 1.05 | 1.70 | 0.0193 |
|  | No conditioning | 52 | 1.59 | 0.57 | 4.43 | 0.3753 |
|  | Missing | 19 | 0.92 | 0.29 | 2.93 | 0.8912 |
| ATG/alemtuzumab | ATG alone | 2070 | 1.00 | . | . | 0.0068 |
|  | No ATG or alemtuzumab | 2092 | 1.27 | 1.06 | 1.53 | 0.0116 |
|  | Missing | 1178 | 1.35 | 1.08 | 1.69 | 0.0078 |
| Graft type | Bone marrow | 3260 | 1.00 | . | . | 0.0078 |
|  | Peripheral blood | 758 | 1.30 | 1.08 | 1.56 | 0.0062 |
|  | Cord blood | 1322 | 1.26 | 0.98 | 1.61 | 0.0682 |
| Donor (BM/PB only) | HLA-matched sib | 1595 | 1.00 | . | . | 0.0271 |
|  | Other related | 322 | 1.09 | 0.80 | 1.47 | 0.5903 |
|  | Well matched URD | 1478 | 1.03 | 0.84 | 1.26 | 0.7819 |
|  | Partially matched URD | 563 | 1.46 | 1.14 | 1.87 | 0.0029 |
|  | URD, match unknown | 60 | 1.35 | 0.71 | 2.56 | 0.3630 |
| Donor-recipient sex match (BM & PB only) | M-M | 1418 | 1.00 | . | . | 0.0035 |
|  | M-F | 875 | 0.87 | 0.69 | 1.09 | 0.2223 |
|  | F-M | 954 | 1.19 | 0.96 | 1.47 | 0.1140 |
|  | F-F | 763 | 1.37 | 1.10 | 1.71 | 0.0049 |
|  | Missing | 8 | 1.19 | 0.28 | 5.02 | 0.8170 |
| Recipient CMV | Negative | 2409 | 1.00 | . | . | 0.0945 |
|  | Positive | 2840 | 1.17 | 1.02 | 1.35 | 0.0299 |
|  | Unknown | 91 | 1.08 | 0.55 | 2.10 | 0.8213 |
| ***Post-transplant*** | | | | | | |
| Time from 1^st^ HCT to PICU | No PICU within one year | 4860 | 1.00 | . | . | <.0001 |
|  | 0-29 days | 175 | 1.19 | 0.80 | 1.78 | 0.3950 |
|  | 30-99 days | 119 | 1.89 | 1.27 | 2.82 | 0.0018 |
|  | 100-365 days | 186 | 2.20 | 1.69 | 2.86 | <.0001 |
| CGVHD before 1 yr | No | 3837 | 1.00 | . | . | <.0001 |
|  | Yes | 1416 | 1.49 | 1.28 | 1.75 | <.0001 |
|  | Missing | 87 | 1.19 | 0.68 | 2.09 | 0.5357 |
| Relapse before 1yr  (malignant disease) | No | 2541 | 1.00 | . | . | <.0001 |
|  | Yes | 276 | 6.80 | 5.60 | 8.25 | <.0001 |
|  | Missing | 53 | 3.34 | 2.27 | 4.92 | <.0001 |

**Legend:** Multivariable model for long-term survival among patients who survived to day +365.

**Supplemental Table 10. Late Effects (CRF subset)**

| **Variable** | **N (%)** |
| --- | --- |
| Congestive heart failure |  |
| No | 2295 (97) |
| Yes | 29 (1) |
| Missing | 47 (2) |
| Diabetes/hyperglycemia |  |
| No | 2067 (87) |
| Yes | 162 (7) |
| Missing | 142 (6) |
| Renal failure severe enough to warrant dialysis |  |
| No | 2218 (94) |
| Yes | 106 (4) |
| Missing | 47 (2) |
| Seizures/Stroke |  |
| No | 2156 (91) |
| Yes | 168 (7) |
| Missing | 47 (2) |
| Non-infectious pulmonary dysfunction |  |
| No | 1897 (80) |
| Yes | 427 (18) |
| Missing | 47 (2) |
| Non-infectious liver toxicity |  |
| No | 1783 (75) |
| Yes | 541 (23) |
| Missing | 47 (2) |

**Legend:** Organ toxicities as reported in the CIBMTR CRF-track.
